# Donor-Specific Antibody Testing is an Effective Surveillance Strategy for High-Risk Antibody Mediated Rejection in Heart Transplant Patients in the Contemporary Era

**DOI:** 10.1101/2023.12.01.23299311

**Authors:** Vincenzo Cusi, Ashley Cardenas, Yuko Tada, Florin Vaida, Nicholas Wettersten, Jennifer Chak, Victor Pretorius, Marcus Anthony Urey, Gerald P. Morris, Grace Lin, Paul J. Kim

**Author notes:** **Address for Correspondence:** Paul J. Kim, MD Division of Cardiovascular Medicine Department of Medicine UC San Diego Health E-mail: pjk017 at health dot ucsd dot edu. **Author contributions:** VC: writing, review and editing, data curation, project administration; AC: conceptualization, writing, data curation; YT: writing, review and editing, data curation, adjudication of clinical outcomes; FV: writing, reviewing, editing, and statistical analyses; NW: writing, review and editing, data curation, adjudication of clinical outcomes; JC: writing, review and editing, data curation; PB: writing, review and editing, data curation; VP: writing, review and editing; MAU: writing, review and editing; GPM: writing, review, and editing, project administration; GL: conceptualization, writing, review and editing, project administration; PJK: conceptualization, writing, review and editing, data curation, adjudication of clinical outcomes, project administration.

## Abstract

**Background:** Pathologic antibody mediated rejection (pAMR) evaluation and donor specific antibody (DSA) testing are recommended in the first year after heart transplantation (HTx) in adult patients. Whether DSA testing adds prognostic information to contemporary pAMR surveillance has not been fully studied.

**Methods:** This was a single center study of consecutive endomyocardial biopsies (EMB) performed between November 2010 and February 2023 in adult HTx patients. The primary objective was to evaluate whether DSA testing contributes additional information to pAMR surveillance to better predict overall survival. Secondary endpoints included cardiac allograft dysfunction and loss.

**Results:** A total of 6,033 EMBs from 544 HTx patients were reviewed for the study. The pAMR+/DSA+ group had significantly lower overall survival versus the pAMR-/DSA-group (hazard ratio [HR] = 2.63; 95% confidence interval [CI], 1.35-5.11; p_c_ = 0.013). In the pAMR+/DSA+ group, patients with cardiac allograft dysfunction, compared to those without allograft dysfunction, had significantly lower overall and cardiac survival (p_c_ < 0.001 for both). In contrast, pAMR+/DSA+ and pAMR-/DSA-patients without cardiac allograft dysfunction showed no difference in overall and cardiac survival. Primary graft dysfunction (PGD) was a novel risk factor for development of *de novo* DSAs (dnDSA) three weeks post-HTx (p = 0.007).

**Conclusions:** DSA testing as the primary surveillance method can identify high-risk pAMR+/DSA+ patients. Surveillance pAMR testing in the contemporary era may need to be reevaluated. Earlier DSA testing at 10-14 days post-HTx should be considered in PGD patients.

## Introduction

Recognition and standardization for the diagnosis of pAMR occurred in 2013 by the International Society for Heart and Lung Transplantation (ISHLT) working group, where surveillance for pAMR in adult HTx patients was first recommended.^1^ With the goal of making pAMR a pathologic diagnosis, akin to acute cellular rejection (ACR),^2^ the ISHLT working group also removed the presence of DSA and cardiac allograft dysfunction for pAMR diagnosis. These pivotal changes were also made to address the concern for underdetection of asymptomatic pAMR.

While DSA testing continues to be recommended with pAMR surveillance in the first year after HTx,^3^ how both results should influence management of HTx patients remains unclear.^4^ In this single-center study, we aimed to evaluate whether DSA testing provides additional information to pAMR surveillance to predict overall survival in HTx patients. In addition, because HTx population demographics and HTx management have significantly changed over time,^5^ we performed a comprehensive analysis to identify potential predictors for dnDSAs, pAMR, cardiac allograft dysfunction, CAV, cardiac and overall survival in patients with pAMR and DSA testing for the contemporary era (2010-current).

## Methods

### Data Sharing

The data that support the findings of this study are openly available in Mendeley Data at 10.17632/d4f7g8hs5z.1.

### Study Design

Consecutive patients who were 18 years of age or older and underwent HTx between November 2010 to February 2023 were retrospectively reviewed. Patients without prior pAMR and DSA results available were excluded. Database lock occurred March 2024, one year after the inclusion of the final patient. The typical EMB surveillance protocol^6^ at the University of California, San Diego Health (UCSD) includes C4d immunofluorescence at 10-14 days post-HTx and subsequently as recommended frequency by the ISHLT.^1^ DSA testing is also performed at the same time intervals.^3^ VC, AC, PB, and JC collected patient data and clinical outcomes from the electronic medical record. Approval for this study was provided by the UCSD Office of IRB Administration (IRB #805675). This study adheres to the principles of the Declaration of Helsinki formulated by the World Medical Association, the Declaration of Istanbul, and the International Society for Heart and Lung Transplantation statement on Transplant Ethics.

### Pathologic Tissue Exams and Anti-Human Leukocyte Antigen (HLA) Antibody Testing

C4d immunofluorescence was performed starting November 2010 with positivity defined according to the ISHLT Working Formulation; however, contemporary ISHLT pAMR grading was implemented at UCSD in July 2015.^1^ Thus, EMB samples prior to July 2015 were regraded using the current pAMR grading scheme for this study (GL). Anti-HLA testing is performed using single-antigen bead LABScreen HLA Class I and II assays (One Lambda, Canoga Park, CA) on LabScan 100 and FlexMap 3D (Luminex, Austin, TX) instruments. Data is analyzed using HLA Fusion software (One Lambda). Antibodies with normalized mean fluorescence intensity (MFI) values > 3,000 are identified as positive, based upon likelihood of causing a positive flow cytometric crossmatch.^7^ DSA are identified by comparison of antibody testing results to donor HLA typing. Concurrent DSA positivity was defined as occurring within a month of a pAMR diagnosis.

### Clinical Outcomes and Variables

The primary outcome was all-cause death or cardiac retransplant. Cause of death was also adjudicated by three experienced cardiologists (NW, YT, PJK).^6^ Secondary outcomes evaluated were: cardiac allograft failure, future episodes of pAMR or dnDSA detection, concurrent or future cardiac allograft dysfunction (echocardiogram demonstrating left ventricular ejection fraction < 50%)^8^ occurring after one week post-HTx, and ISHLT cardiac allograft vasculopathy (CAV) grade 2 or greater^9^. PGD diagnosis was based on documentation by the HTx clinical team or need of extracorporeal membrane oxygenation and/or percutaneous mechanical circulatory support after HTx.^10^ Documentation by a clinical team member of immunosuppressive medication nonadherence after HTx was recorded retrospectively by medical chart review and independent of the study (VC, AC, PB, JC).^11–13^

### Statistical Analysis

Demographic and clinical variables were analyzed with standard statistics as previously described for continuous and count variables.^6^ The association of pAMR/DSA groups with time to event outcomes was evaluated using single predictor and multipredictor Cox proportional hazards models. The multipredictor models for overall and cardiac survival were adjusted for recipient age, sex, and race/ethnicity. Additional exploratory analyses investigated factors associated with time to event outcomes using Cox models applying a forward model selection procedure with p-value ≤ 0.15 threshold for inclusion. Cox models with time-dependent covariates were used when the proportional hazards assumption of constant hazard ratios was violated. To adjust for different causes of death as competing outcomes, a competing-risk regression model was constructed using the Fine and Gray method. We implemented bootstrapping, repeated 10,000 times, to generate approximate sampling distributions for the statistics of interest. The mean and 95% CI for each statistic were taken from the bootstrap sampling distribution.

Analysis was conducted in R (R Core Team, 2022). We used the Bonferroni-Holm procedure whenever multiple comparisons were performed while implementing a particular statistical hypothesis test. The corrected p-values are designated as p_c_. For single hypothesis testing, we report the uncorrected p-value. P or p_c_ < 0.05 are considered significant.

## Results

### Patient Demographics

A total of 6,033 EMBs from 544 HTx patients, including four cardiac retransplants, were evaluated (**Figure 1**). We divided all patients into one of four groups based on history of pAMR and DSA results: pAMR+/DSA+ (n=45, 8.3%), pAMR+/DSA- (n=30, 5.5%), pAMR-/DSA+ (n=95, 17.5%), and pAMR-/DSA- (n=374, 68.8%) patients.

**Figure 1.**
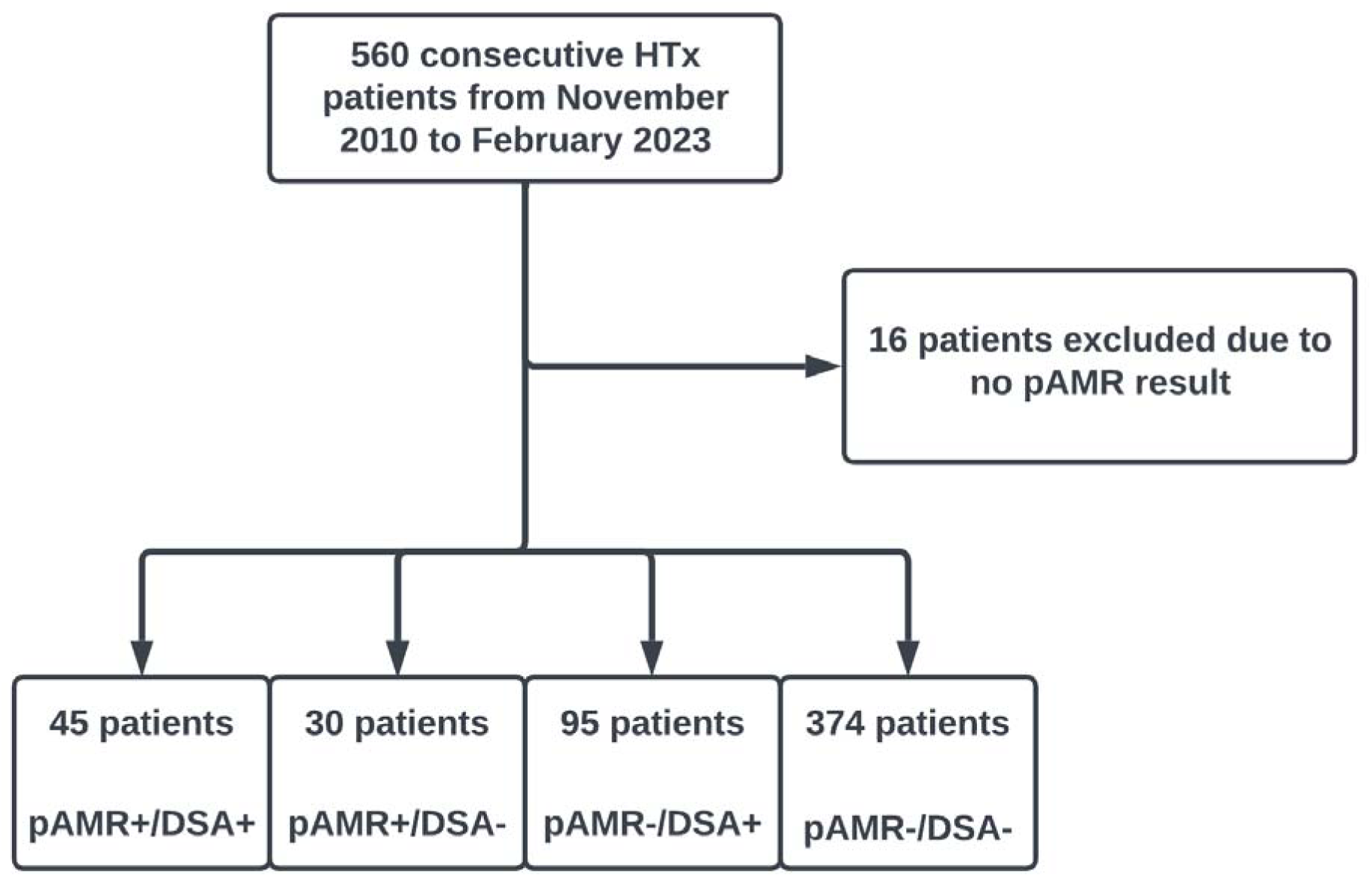
Flow diagram for heart transplants included then grouped based on history of pAMR and DSA results. HTx, heart transplantation; DSA, donor-specific antibodies; pAMR, pathologic antibody mediated rejection.

Characteristics of the study population are summarized in **Tables 1** and **S1**. HTx recipients were followed for a total of 1,999.2 person-years from the time of HTx.

**Table 1.**
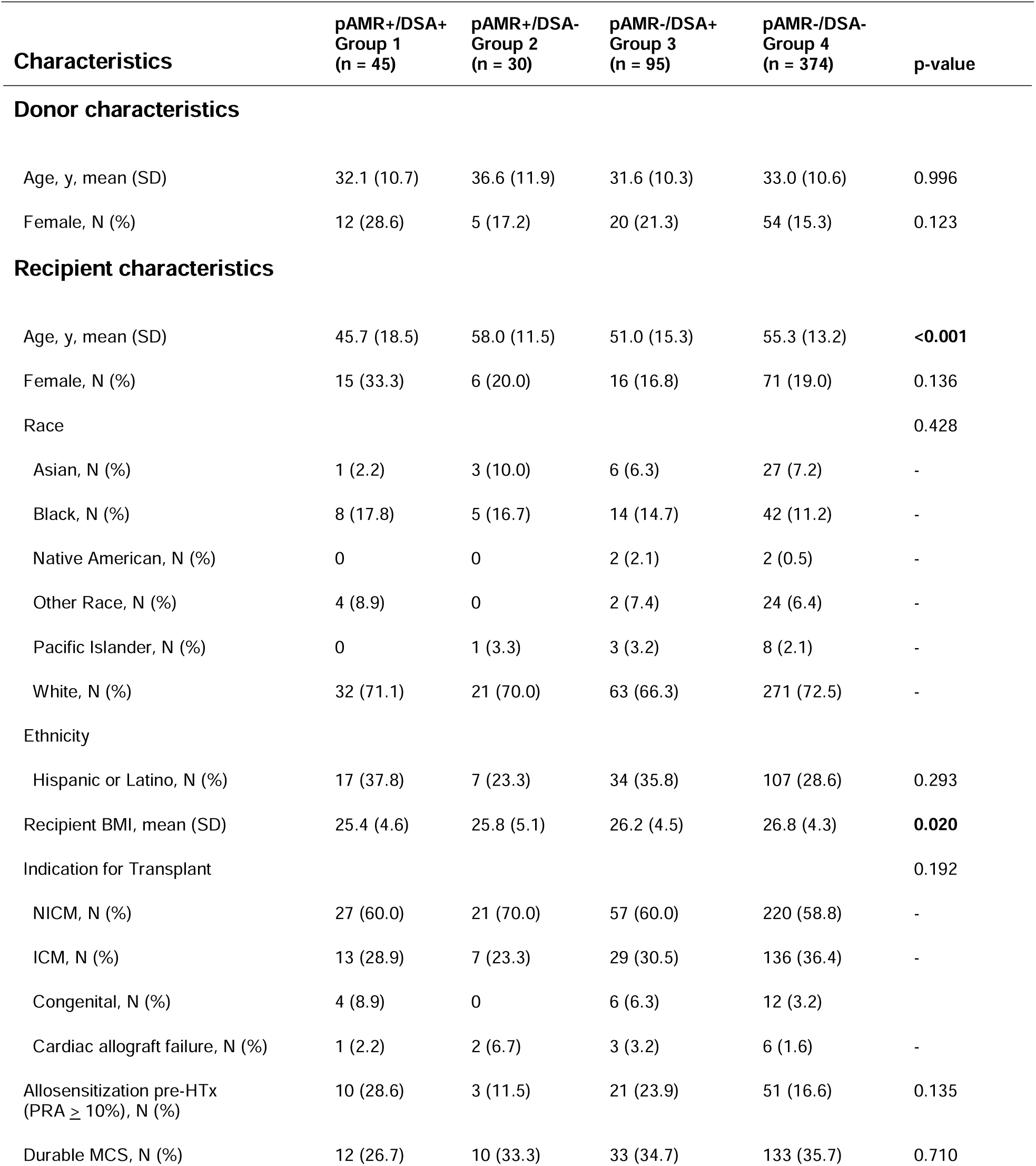
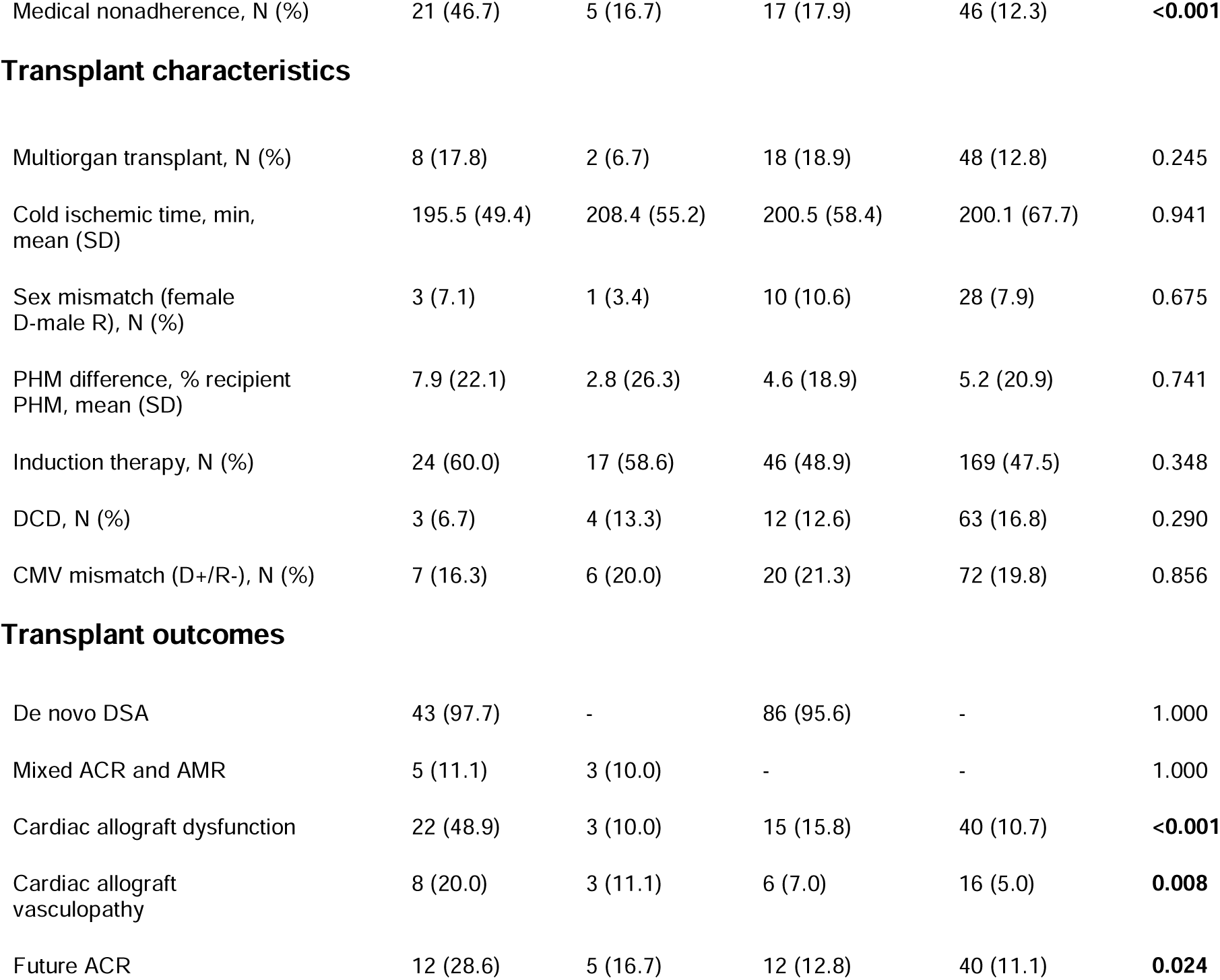
Baseline characteristics of heart transplant patients by pAMR and DSA results. ACR, acute cellular rejection; AMR, antibody mediated rejection; BMI, body mass index; CMV, cytomegalovirus; DCD, donation after cardiac death; DSA, donor-specific antibodies; HTx, heart transplantation; ICM, ischemic cardiomyopathy; MCS, mechanical circulatory support; NICM, nonischemic cardiomyopathy; PHM, predicted heart mass; PRA, panel reactive antibodies.

| Characteristics | pAMR+/DSA+<br>Group 1<br>(n = 45) | pAMR+/DSA-<br>Group 2<br>(n = 30) | pAMR-/DSA+<br>Group 3<br>(n = 95) | pAMR-/DSA-<br>Group 4<br>(n = 374) | p-value |
| --- | --- | --- | --- | --- | --- |
| **Donor characteristics** |  |  |  |  |  |
| Age, y, mean (SD) | 32.1 (10.7) | 36.6 (11.9) | 31.6 (10.3) | 33.0 (10.6) | 0.996 |
| Female, N (%) | 12 (28.6) | 5 (17.2) | 20 (21.3) | 54 (15.3) | 0.123 |
| **Recipient characteristics** |  |  |  |  |  |
| Age, y, mean (SD) | 45.7 (18.5) | 58.0 (11.5) | 51.0 (15.3) | 55.3 (13.2) | **<0.001** |
| Female, N (%) | 15 (33.3) | 6 (20.0) | 16 (16.8) | 71 (19.0) | 0.136 |
| Race |  |  |  |  | 0.428 |
| Asian, N (%) | 1 (2.2) | 3 (10.0) | 6 (6.3) | 27 (7.2) | - |
| Black, N (%) | 8 (17.8) | 5 (16.7) | 14 (14.7) | 42 (11.2) | - |
| Native American, N (%) | 0 | 0 | 2 (2.1) | 2 (0.5) | - |
| Other Race, N (%) | 4 (8.9) | 0 | 2 (7.4) | 24 (6.4) | - |
| Pacific Islander, N (%) | 0 | 1 (3.3) | 3 (3.2) | 8 (2.1) | - |
| White, N (%) | 32 (71.1) | 21 (70.0) | 63 (66.3) | 271 (72.5) | - |
| Ethnicity |  |  |  |  |  |
| Hispanic or Latino, N (%) | 17 (37.8) | 7 (23.3) | 34 (35.8) | 107 (28.6) | 0.293 |
| Recipient BMI, mean (SD) | 25.4 (4.6) | 25.8 (5.1) | 26.2 (4.5) | 26.8 (4.3) | **0.020** |
| Indication for Transplant |  |  |  |  | 0.192 |
| NICM, N (%) | 27 (60.0) | 21 (70.0) | 57 (60.0) | 220 (58.8) | - |
| ICM, N (%) | 13 (28.9) | 7 (23.3) | 29 (30.5) | 136 (36.4) | - |
| Congenital, N (%) | 4 (8.9) | 0 | 6 (6.3) | 12 (3.2) | - |
| Cardiac allograft failure, N (%) | 1 (2.2) | 2 (6.7) | 3 (3.2) | 6 (1.6) | - |
| Allosensitization pre-HTx<br>(PRA ≥ 10%), N (%) | 10 (28.6) | 3 (11.5) | 21 (23.9) | 51 (16.6) | 0.135 |
| Durable MCS, N (%) | 12 (26.7) | 10 (33.3) | 33 (34.7) | 133 (35.7) | 0.710 |
| Medical nonadherence, N (%) | 21 (46.7) | 5 (16.7) | 17 (17.9) | 46 (12.3) | **<0.001** |
| Transplant characteristics |  |  |  |  |  |
| Multiorgan transplant, N (%) | 8 (17.8) | 2 (6.7) | 18 (18.9) | 48 (12.8) | 0.245 |
| Cold ischemic time, min, mean (SD) | 195.5 (49.4) | 208.4 (55.2) | 200.5 (58.4) | 200.1 (67.7) | 0.941 |
| Sex mismatch (female D-male R), N (%) | 3 (7.1) | 1 (3.4) | 10 (10.6) | 28 (7.9) | 0.675 |
| PHM difference, % recipient PHM, mean (SD) | 7.9 (22.1) | 2.8 (26.3) | 4.6 (18.9) | 5.2 (20.9) | 0.741 |
| Induction therapy, N (%) | 24 (60.0) | 17 (58.6) | 46 (48.9) | 169 (47.5) | 0.348 |
| DCD, N (%) | 3 (6.7) | 4 (13.3) | 12 (12.6) | 63 (16.8) | 0.290 |
| CMV mismatch (D+/R-), N (%) | 7 (16.3) | 6 (20.0) | 20 (21.3) | 72 (19.8) | 0.856 |
| Transplant outcomes |  |  |  |  |  |
| De novo DSA | 43 (97.7) | - | 86 (95.6) | - | 1.000 |
| Mixed ACR and AMR | 5 (11.1) | 3 (10.0) | - | - | 1.000 |
| Cardiac allograft dysfunction | 22 (48.9) | 3 (10.0) | 15 (15.8) | 40 (10.7) | **<0.001** |
| Cardiac allograft vasculopathy | 8 (20.0) | 3 (11.1) | 6 (7.0) | 16 (5.0) | **0.008** |
| Future ACR | 12 (28.6) | 5 (16.7) | 12 (12.8) | 40 (11.1) | **0.024** |

### Association of pAMR/DSA classification with overall and cardiac survival

A total of 61 (11.2%) patients died or underwent cardiac retransplant during the follow-up period. Adjudicated causes of death are provided in **Table S2**. Initial adjudication of cause of death agreed 87.7% of the time with a Cohen’s kappa of 0.82 (0.69, 0.94; p < 0.001).

Overall survival was significantly lower in pAMR+/DSA+ compared to pAMR-/DSA-patients (HR = 2.63; 95% CI, 1.35-5.11; p_c_ = 0.013; **Figure 2A**). Cardiac survival was also significantly lower in the pAMR+/DSA+ compared to pAMR-/DSA-group (HR = 7.00; 95% CI, 2.31-21.20; p_c_ = 0.002; **Figure 2B**). There was no significant difference in overall or cardiac survival in the pAMR+/DSA- (p_c_ = 1.000) and pAMR-/DSA+ groups (p_c_ = 1.000) compared to pAMR-/DSA-patients. Sensitivity analyses performed with dnDSAs and initial pAMR/DSA classification from the first pAMR+ or DSA+ result, with the concurrent corresponding DSA or pAMR test result, also demonstrated similar findings (not shown).

**Figure 2.**
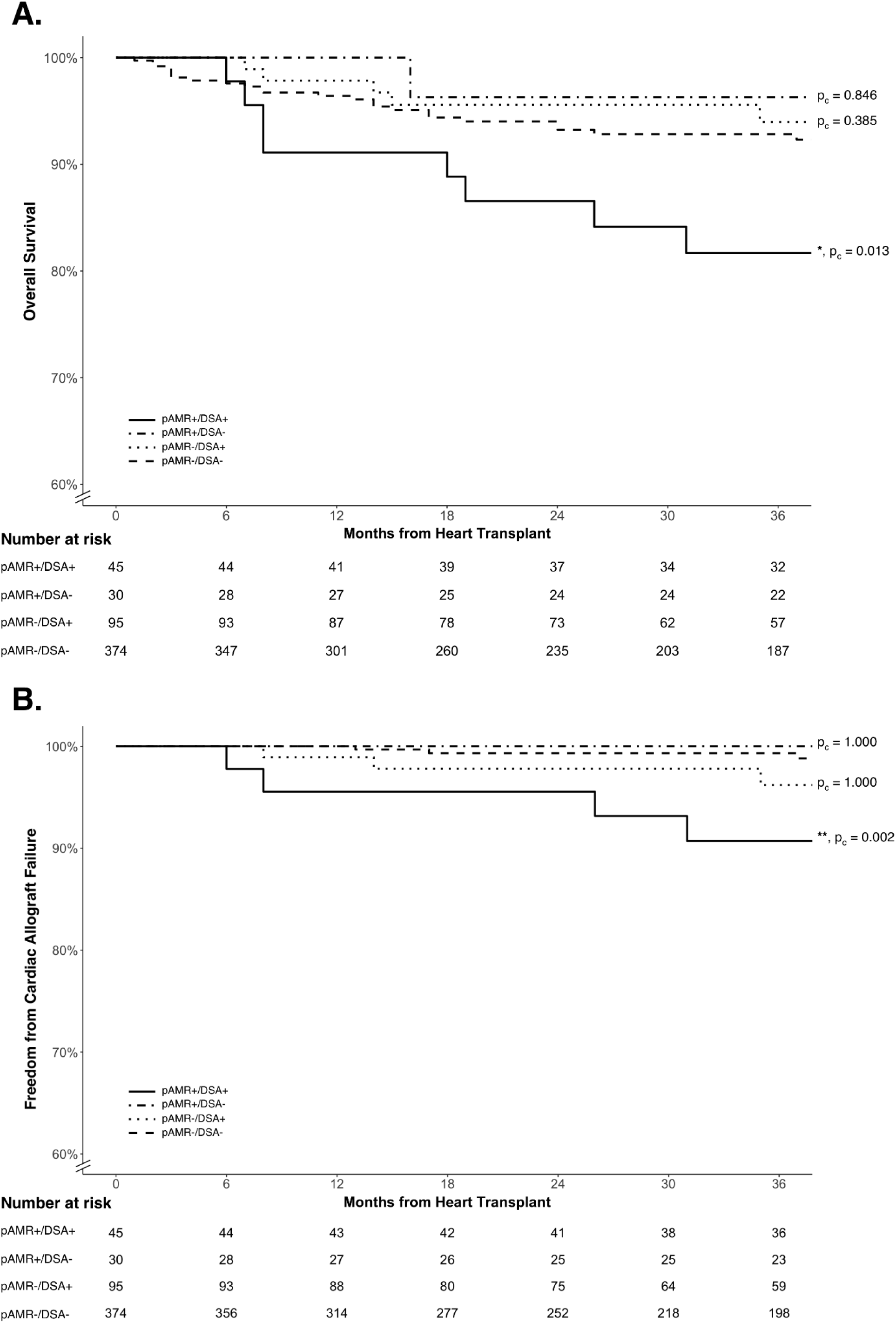
Kaplan-Meier Curves by pAMR/DSA Status for (**A**) Overall and (**B**) Cardiac Survival. The pAMR+/DSA+ patients show significantly reduced overall and cardiac survival compared to the pAMR-/DSA-group. The pAMR+/DSA- and pAMR-/DSA+ groups were not significantly different from the pAMR-/DSA-group. Fine-Gray subdistribution hazard model was used to account for competing causes of death. Adjusted p-values for pairwise comparisons compared to the pAMR-/DSA-reference group are provided next to the survival curves. DSA, donor-specific antibodies; pAMR, pathologic antibody mediated rejection.

### Analysis of pAMR+/DSA+ patients

The pAMR+/DSA+ patients demonstrated significantly later diagnosis of pAMR after HTx compared to the pAMR+/DSA-patients (33.6 vs 3.7 weeks; p = 0.004). The majority (62.2%) of pAMR+/DSA+ patients were diagnosed by pAMR+ and DSA+ results concurrently, i.e., within one month of either test. A small number of patients (6.7%) were found to have a DSA+ results three months after a pAMR+ result. No patient with an initial pAMR+/DSA-classification had a primary outcome of death or retransplant within a year of a pAMR diagnosis.

There was no significant difference in time to dnDSA positivity in pAMR+/DSA+ compared to pAMR-/DSA+ patients (20.3 vs 17.0 weeks; p = 0.941). Detection of both class I and II dnDSAs on initial DSA+ testing demonstrated the highest positive predictive value than other DSAs, as summarized in **Table S3**. Detection of both class I and II dnDSAs on initial DSA+ testing was also associated with greater odds for pAMR2 or pAMR3 grades than class II dnDSAs alone (OR = 5.92; 95% CI, 1.02-33.74; p_c_ = 0.025) and showed a trend for greater odds compared to class I dnDSAs alone (OR = 4.67; 95% CI, 0.70-36.86; p_c_ = 0.147). Detection of both class I and II dnDSAs at any time post-HTx had a significantly higher risk of pAMR, cardiac allograft dysfunction, CAV, cardiac allograft failure, and all-cause death or retransplant (p < 0.001 for all). Class II dnDSAs alone (p < 0.001) predicted pAMR but not other clinical outcomes. We did not find specific class II dnDSAs alone that significantly increased the risk for pAMR compared to other class II dnDSAs. Class I dnDSAs alone did not predict pAMR nor other clinical outcomes.

### Cardiac allograft dysfunction in pAMR+/DSA+ patients

We found that a pAMR+/DSA+ status independently predicted cardiac allograft dysfunction (**Table 2**). In contrast, the rates of cardiac allograft dysfunction for pAMR+/DSA- (p = 0.601) and pAMR-/DSA+ groups (p = 0.235) were not significantly different from the pAMR-/DSA-patients, with these three groups demonstrating an occurrence of cardiac allograft dysfunction within the range of 10 to 16%. In pAMR+/DSA+ patients, diagnosis of pAMR in the first year or after 1-year post-HTx also did not show differences in cardiac allograft dysfunction, cardiac survival, or overall survival (not shown).

**Table 2.** Unipredictor and multipredictor Cox proportional hazards analyses for cardiac allograft dysfunction. Unipredictor parameters with a p-value ≤ 0.15 are included in addition to certain clinical parameters of interest. HRs and CIs are not provided for categorical variables in this table. ACR, acute cellular rejection; CI, confidence interval; cPRA, calculated panel reactive antibodies; DSA, donor-specific antibodies; ECMO, extracorporeal membrane oxygenation; HR, hazard ratio; HTx, heart transplantation; MCS, mechanical circulatory support; pAMR, pathologic antibody mediated rejection; PHM, predicted heart mass; pMCS, percutaneous mechanical circulatory support; UNOS, United Network for Organ Sharing. *, allosensitized patients defined as having a UNOS cPRA >= 10%.

| Predictor | No. of patients<br>(n = 544) | No. of events<br>(n = 80) | HR | 95% CI | p-value |
| --- | --- | --- | --- | --- | --- |
| **Unipredictor analysis** |  |  |  |  |  |
| HTx indication (vs. non-ischemic cardiomyopathy) | 544 | 80 | - | - | **0.020** |
| Allosensitization pre-HTx* | 457 | 67 | 0.89 | 0.48-1.67 | 0.715 |
| Durable MCS at time of HTx (yes vs. no) | 543 | 80 | 1.44 | 0.92-2.24 | 0.107 |
| Medical nonadherence (yes vs. no) | 544 | 80 | 2.64 | 1.67-4.18 | **<0.001** |
| Donor age (by 10-yr) | 520 | 78 | 0.88 | 0.71-1.08 | 0.207 |
| Induction therapy (yes vs. no) | 519 | 77 | 0.84 | 0.53-1.33 | 0.458 |
| Cold ischemic time (per hour) | 519 | 78 | 0.90 | 0.73-1.11 | 0.330 |
| PHM difference (per % recipient PHM increment) | 513 | 77 | 1.01 | 1.00-1.02 | 0.130 |
| Donation after cardiac death (vs. brain death) | 544 | 80 | 1.37 | 0.69-2.71 | 0.373 |
| ECMO pre-HTx (yes vs. no) | 540 | 79 | 2.62 | 0.82-8.32 | 0.103 |
| pMCS pre-HTx (yes vs. no) | 540 | 79 | 0.55 | 0.28-1.11 | 0.096 |
| Primary graft dysfunction (yes vs. no) | 542 | 80 | 2.95 | 1.87-4.67 | **<0.001** |
| De novo DSAs (vs. no DSA) | 528 | 73 | 2.17 | 1.35-3.46 | **0.001** |
| Class I de novo DSAs alone (vs. no DSA) | 528 | 73 | 0.40 | 0.06-2.91 | 0.366 |
| Class II de novo DSAs alone (vs. no DSA) | 528 | 73 | 1.41 | 0.74-2.69 | 0.292 |
| Both class I and II de novo DSAs (vs. no DSA) | 528 | 73 | 5.91 | 3.36-10.40 | **<0.001** |
| Concurrent ACR grade with pAMR+ diagnosis (vs. pAMR+ with ACR grade 0R) | 75 | 25 | - | - | 0.301 |
| CMV mismatch (vs. CMV D-/R-) | 531 | 80 | - | - | 0.190 |
| Sex mismatch (female D-male R vs. male D-male R) | 519 | 78 | 0.58 | 0.21-1.58 | 0.285 |
| pAMR/DSA group (vs. pAMR-/DSA- group) | 544 | 80 | - | - | **<0.001** |
| Cardiac allograft Vasculopathy (vs. CAV grades 0 or 1) | 445 | 63 | 8.82 | 3.18-24.44 | **<0.001** |
| History of ACR > 1R (vs. ACR grades 0R/1R) | 514 | 76 | 1.54 | 0.88-2.72 | 0.133 |
| **Multipredictor analysis** |  |  |  |  |  |
| Medical nonadherence | 544 | 80 | 2.36 | 1.36-4.08 | **0.002** |
| Primary graft dysfunction | 542 | 80 | 2.25 | 1.29-3.92 | **0.004** |
| Ischemic cardiomyopathy as HTx indication* | 544 | 80 | 2.07 | 1.22-3.51 | **0.007** |
| pAMR+/DSA+ group<sup>#</sup> | 544 | 80 | 2.36 | 1.19-4.66 | **0.014** |
| Cardiac allograft vasculopathy | 445 | 63 | 3.67 | 1.21-11.10 | **0.021** |
| pMCS pre-HTx | 540 | 79 | 0.46 | 0.19-1.11 | 0.083 |
| PHM difference | 513 | 77 | 1.01 | 1.00-1.02 | 0.084 |

Presence of cardiac allograft dysfunction was associated with lower overall and cardiac survival (**Table 3** and **Table S4**). In the pAMR+/DSA+ group, patients with cardiac allograft dysfunction had significantly lower overall and cardiac survival compared to those without allograft dysfunction (**Figure 3**). Thus, lower overall and cardiac survival in pAMR+/DSA+ patients were mediated by cardiac allograft dysfunction.

**Table 3.** Unipredictor and multipredictor Cox proportional hazards analyses for overall survival. Unipredictor parameters with a p-value ≤ 0.15 are included in addition to certain clinical parameters of interest. HRs and CIs are not provided for categorical variables in this table. ACR, acute cellular rejection; CI, confidence interval; CMV, cytomegalovirus; cPRA, calculated panel reactive antibodies; DSA, donor-specific antibodies; ECMO, extracorporeal membrane oxygenation; HTx, heart transplantation; HR, hazard ratio; MCS, mechanical circulatory support; pAMR, pathologic antibody mediated rejection; PHM, predicted heart mass; pMCS, percutaneous mechanical circulatory support; UNOS, United Network for Organ Sharing. *, allosensitized patients defined as having a UNOS cPRA >= 10%.

| Predictor | No. of patients<br>(n = 544) | No. of events<br>(n = 61) | HR | 95% CI | p-value |
| --- | --- | --- | --- | --- | --- |
| **Unipredictor analysis** |  |  |  |  |  |
| Recipient age (by 10-yr) | 544 | 61 | 1.02 | 0.86-1.21 | 0.793 |
| Recipient race and ethnicity (vs. non-Hispanic White) | 544 | 61 | - | - | **0.027** |
| Multiorgan Transplant (yes vs. no) | 544 | 61 | 0.62 | 0.25-1.55 | 0.304 |
| HTx indication (vs. non-ischemic cardiomyopathy) | 544 | 61 | - | - | 0.403 |
| Allosensitization pre-HTx* | 457 | 44 | 0.98 | 0.47-2.05 | 0.962 |
| Durable MCS at time of HTx (yes vs. no) | 543 | 61 | 1.13 | 0.68-1.89 | 0.639 |
| Medical nonadherence (yes vs. no) | 544 | 61 | 2.33 | 1.37-3.96 | **0.002** |
| Donor age (by 10-yr) | 520 | 58 | 0.97 | 0.77-1.23 | 0.814 |
| Induction therapy (yes vs. no) | 519 | 58 | 1.23 | 0.71-2.11 | 0.463 |
| Cold ischemic time (per hour) | 519 | 58 | 0.86 | 0.66-1.11 | 0.238 |
| PHM difference (per % recipient PHM increment) | 513 | 58 | 1.00 | 0.99-1.01 | 0.775 |
| Donation after cardiac death (vs. brain death) | 544 | 61 | 1.16 | 0.45-3.01 | 0.759 |
| ECMO pre-HTx (yes vs. no) | 540 | 60 | 2.34 | 0.57-9.63 | 0.237 |
| pMCS pre-HTx (yes vs. no) | 540 | 60 | 1.37 | 0.72-2.61 | 0.337 |
| Primary graft dysfunction (yes vs. no) | 542 | 61 | 2.51 | 1.46-4.32 | **0.001** |
| De novo DSAs (vs. no DSAs) | 533 | 59 | 1.19 | 0.67-2.10 | 0.550 |
| Class I de novo DSAs alone (vs. no DSAs) | 533 | 59 | - | - | 0.996 |
| Class II de novo DSAs alone (vs. no DSAs) | 533 | 59 | 0.95 | 0.45-2.03 | 0.897 |
| Both class I and II de novo DSAs (vs. no DSAs) | 533 | 59 | 2.64 | 1.28-5.44 | **0.009** |
| CMV mismatch (vs. D-/R-) | 531 | 60 | - | - | 0.593 |
| Sex mismatch (female D-male R vs. male D-male R) | 519 | 58 | 2.27 | 1.16-4.42 | **0.016** |
| pAMR/DSA group (vs. pAMR-/DSA- group) | 544 | 61 | - | - | **0.018** |
| Cardiac allograft vasculopathy (vs. CAV grades 0 or 1) | 474 | 41 | 2.48 | 1.17-5.24 | **0.018** |
| Cardiac allograft dysfunction (yes vs. no) | 544 | 61 | 2.74 | 1.61-4.66 | **<0.001** |
| History of ACR > 1R (vs. ACR grades 0R/1R) | 530 | 61 | 1.56 | 0.88-2.76 | 0.130 |
| **Multipredictor analysis** |  |  |  |  |  |
| Cardiac allograft dysfunction | 544 | 61 | 2.27 | 1.27-4.06 | **0.006** |
| Medical nonadherence | 544 | 61 | 1.93 | 1.09-3.39 | **0.023** |
| Primary graft dysfunction | 542 | 61 | 2.09 | 1.18-3.71 | **0.012** |
| Recipient race and ethnicity | 544 | 61 | - | - | **0.035** |

**Figure 3.**
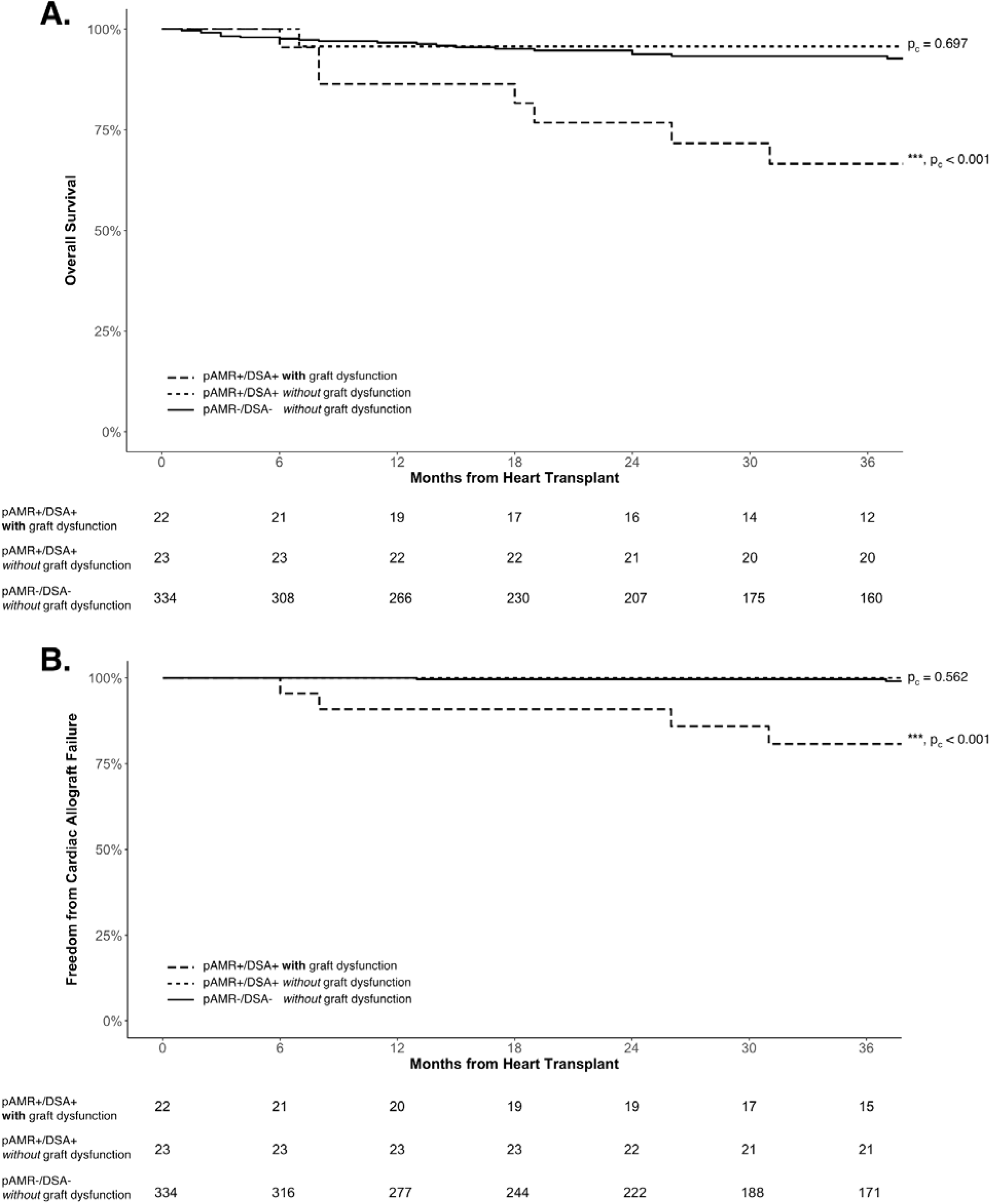
Kaplan-Meier Curves by Cardiac Allograft Dysfunction for (**A**) Overall and (**B**) Cardiac Survival. The pAMR+/DSA+ patients with graft dysfunction showed significantly worse overall and cardiac survival compared to both pAMR+/DSA+ and pAMR-/DSA-patients without graft dysfunction. Adjusted p-values for pairwise comparisons compared to the pAMR-/DSA-patients without allograft dysfunction, as a reference group, are provided next to the survival curves. DSA, donor-specific antibodies; pAMR, pathologic antibody mediated rejection.

### Predictors for dnDSAs

Younger recipient age, medication nonadherence, and PGD were independent predictors for development of dnDSAs (**Table 4**). Patients with PGD demonstrated a significant increase in dnDSAs within three weeks post-HTx (**Figure 4**). PGD also showed a trend towards increased pAMR (HR = 1.66; 95% CI, 0.98-2.79; p = 0.058). Additional subgroup analyses did not show utilization of extracorporeal membrane oxygenation post-HTx or donation after circulatory death (DCD) to be significantly associated with dnDSAs.

**Table 4.** Unipredictor and multipredictor Cox proportional hazards analyses for de novo donor-specific antibody positivity. Unipredictor parameters with a p-value ≤ 0.15 are included in addition to certain clinical parameters of interest. HRs and CIs are not provided for categorical variables in this table. ACR, acute cellular rejection; CI, confidence interval; CMV, cytomegalovirus; cPRA, calculated panel reactive antibodies; DSA, donor-specific antibodies; ECMO, extracorporeal membrane oxygenation; HTx, heart transplantation; HR, hazard ratio; MCS, mechanical circulatory support; pAMR, pathologic antibody mediated rejection; pMCS, percutaneous mechanical circulatory support; UNOS, United Network for Organ Sharing. *, allosensitized patients defined as having a UNOS cPRA >= 10%.

| Predictor | No. of patients<br>(n = 533) | Total # of events<br>(n = 129) | HR | 95% CI | p-value |
| --- | --- | --- | --- | --- | --- |
| **Unipredictor analysis** |  |  |  |  |  |
| Recipient age (by 10-yr) | 533 | 129 | 0.83 | 0.75-0.93 | **0.001** |
| Recipient race and ethnicity (vs. non-Hispanic White) | 533 | 129 | - | - | 0.127 |
| Multiorgan transplant (yes vs. no) | 533 | 129 | 1.61 | 1.04-2.52 | **0.035** |
| HTx indication (vs. non-ischemic cardiomyopathy) | 533 | 129 | - | - | 0.318 |
| Allosensitization pre-HTx* | 452 | 118 | 1.45 | 0.95-2.22 | 0.084 |
| Blood type (vs. blood type O) | 518 | 129 | - | - | **0.013** |
| Durable MCS at time of HTx (yes vs. no) | 532 | 129 | 0.83 | 0.57-1.19 | 0.310 |
| Medical nonadherence (yes vs. no) | 533 | 129 | 1.87 | 1.27-2.75 | **0.001** |
| Donor age (by 10-yr) | 511 | 127 | 0.89 | 0.76-1.05 | 0.159 |
| Induction therapy (yes vs. no) | 511 | 126 | 1.01 | 0.71-1.44 | 0.947 |
| Cold ischemic time (per hour) | 510 | 126 | 1.03 | 0.87-1.21 | 0.752 |
| Donation after cardiac death (vs. brain death) | 533 | 129 | 0.98 | 0.57-1.69 | 0.936 |
| ECMO pre-HTx (yes vs. no) | 530 | 127 | 2.34 | 0.86-6.35 | 0.095 |
| pMCS pre-HTx (yes vs. no) | 530 | 127 | 0.99 | 0.64-1.54 | 0.974 |
| ECMO post-HTx (yes vs. no) | 530 | 127 | 1.71 | 0.80-3.67 | 0.168 |
| Primary graft dysfunction (yes vs. no) | 531 | 129 | 1.75 | 1.14-2.68 | **0.010** |
| CMV mismatch (vs. D-/R-) | 523 | 129 | - | - | 0.535 |
| Sex mismatch (vs. male D-male R) | 510 | 127 | - | - | 0.701 |
| History of ACR > 1R (vs. ACR grades 0R/1R) | 520 | 129 | 0.56 | 0.31-1.02 | **0.058** |
| **Multipredictor analysis** |  |  |  |  |  |
| Recipient age | 533 | 129 | 0.82 | 0.73-0.91 | **<0.001** |
| Medical nonadherence | 533 | 129 | 1.71 | 1.15-2.52 | **0.007** |
| Primary graft dysfunction | 531 | 129 | 1.67 | 1.13-2.48 | **0.010** |
| Multiorgan transplant | 533 | 129 | 1.49 | 0.95-2.33 | 0.084 |
| History of ACR > 1R | 520 | 129 | 0.60 | 0.33-1.10 | 0.098 |

**Figure 4.**
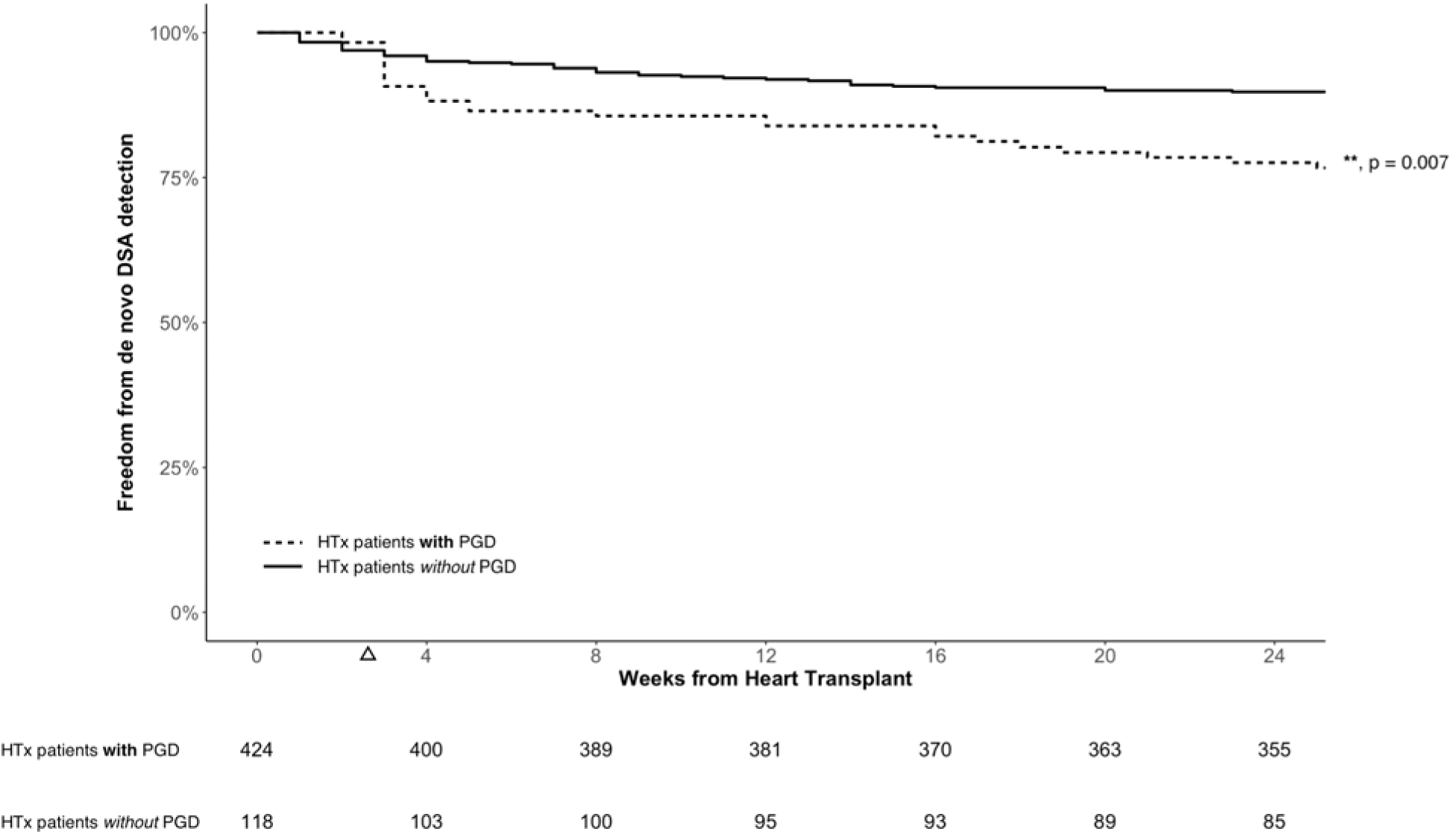
Kaplan-Meier Curves for Freedom from De Novo DSA Detection in HTx patients with and without PGD. HTx patients with PGD demonstrated a significant increase in de novo DSAs three weeks post-HTx (triangle), after which the hazard ratios were not significantly different between HTx patients with and without PGD. DSA, donor-specific antibodies; pAMR, pathologic antibody mediated rejection.

## Discussion

In this retrospective cohort of 544 adult HTx patients with 2,000 patient-years of follow-up, we observed the following key findings. First, DSA testing with contemporary ISHLT pAMR grading identified pAMR+/DSA+ patients to have significantly lower overall and cardiac survival compared to pAMR-/DSA-patients. Second, in the pAMR+/DSA+ group, we found that lower overall and cardiac survival was mediated by cardiac allograft dysfunction. Third, detection of both class I and II dnDSAs had the highest predictive value for pAMR. Fourth, PGD is a novel risk factor for the development of dnDSAs.

With the reevaluation of the utility of surveillance EMBs in the contemporary era,^6^ the current study findings support the use of DSA testing as the primary method of surveillance for clinically relevant pAMR. While Clerkin et al. previously showed no difference in cardiac allograft failure in patients with pAMR or DSAs,^14^ substudy analysis showed that dnDSAs were associated with significantly lower cardiac survival, which are consistent with our findings as well as others.^15,16^ However, we go further to demonstrate that the risk of lower overall and cardiac survival is found specifically in the pAMR+/DSA+ group. Thus, our proposed strategy would redefine the current role of pAMR surveillance^3^ to for-cause testing upon a DSA+ result.^6^ Coutance et al. also recently proposed a clinical prediction model which includes history of a prior ISHLT pAMR2 diagnosis, cardiac allograft dysfunction, and DSA as 3 of the 5 predictor variables.^8^ In contrast, our suggested strategy only requires DSA screening and, more importantly, provides an opportunity to prevent cardiac allograft dysfunction in pAMR+/DSA+ patients. Additionally, we note that new noninvasive biomarker testing, including donor-derived cell-free DNA,^17^ may play a future role in pAMR risk stratification. However, DSA testing will continue to be an essential assay in the foreseeable future and ongoing randomized controlled trials (e.g., NCT06414603: ACES-EMB) are evaluating novel noninvasive biomarkers as an adjunct to DSA testing.

We hypothesize that ischemia-reperfusion tissue injury post-HTx predominantly contributes to early positive C4d immunostaining in pAMR+/DSA-patients, while complement activation due to dnDSAs is likely responsible for later positive C4d immunostaining that occurs in pAMR+/DSA+ patients. Previous studies have shown positive C4d immunostaining in the early post-HTx period could result from the lectin complement pathway related to ischemia-reperfusion tissue injury.^18,19^ Mantell et al. also have shown transcriptomic differences between the pAMR+/DSA+ and pAMR+/DSA-groups, with significant upregulation of genes related to immunity in the pAMR+/DSA+ patients.^20^ In addition, while non-HLA antibodies continue to be investigated in pAMR,^21^ there was no appreciable difference in outcomes in the pAMR+/DSA-compared to the pAMR-/DSA-group to suggest a meaningful clinical effect by non-HLA antibodies.

Furthermore, we found that the pAMR+/DSA+ patients with cardiac allograft dysfunction carried most of the increased risk for lower overall and cardiac survival and accounted for 18% of deaths or cardiac retransplants while representing only 4% of our HTx cohort. Cardiac allograft dysfunction was previously part of the clinical criteria for diagnosis of antibody mediated rejection,^22^ and prior studies have also demonstrated the prognostic importance of cardiac allograft dysfunction, although not specifically in pAMR+/DSA+ patients.^23,24^ Thus, our study highlights the importance of surveillance for cardiac allograft dysfunction in pAMR+/DSA+ patients, given the significant increase in mortality once cardiac allograft dysfunction occurs.^25^ Of note, cardiac allograft dysfunction also occurred in pAMR-/DSA+ and pAMR-/DSA-patients, albeit at much lower rates. The cause was not identified in many cases (36%) and thus were considered to be nonspecific graft dysfunction.^26^

Detection of both class I and II dnDSAs increased the risk for pAMR+ diagnosis three times compared to detection of other dnDSAs. In contrast to some other studies,^14,27^ we were not able to identify specific class II dnDSAs alone that significantly increased the risk for future pAMR more than other class II dnDSAs. We hypothesize some of the differences in literature are related to how HLA-DQ DSAs, the most frequently detected of the DSAs, are categorized when occurring in the presence of other DSAs.^28^ In the current study, we evaluated class I alone, class II alone, and both class I and II dnDSAs as separate categories and also evaluated for progression from one dnDSA category to another in subsequent testing.

Additionally, our study findings indirectly suggest increased immunogenicity associated with detection of both class I and II dnDSAs compared to either class I or II dnDSAs alone. HTx patients with both class I and II dnDSAs demonstrated a higher rate of more severe initial pAMR+ grades compared to patients with either class I or II dnDSAs alone. Patients with both class I and II dnDSAs were also at a significantly increased risk for pAMR-mediated clinical outcomes, including CAV, while patients with either class I or II dnDSAs alone were not. Prior studies have also shown that detection of class I and II DSAs, using the contemporary solid phase assays, were more predictive of persistent and cytotoxic DSAs than either class I or II DSAs alone.^29,30^ While class II DSAs can activate endothelial cells toward a proinflammatory response,^31^ the potential synergistic interaction of both class I and II DSAs warrants further study.

Finally, our data demonstrates PGD as a possible risk factor for development of dnDSAs, providing a novel insight into the potential relationship of PGD, dnDSAs, and pAMR in the peri-transplant period. Han et al. previously showed a similar incidence of dnDSAs in patients with PGD compared to those without PGD.^32^ However, this study included a patient cohort with a much higher preformed DSA prevalence and lower rate of moderate or severe PGD than typically described from other centers,^33^ which likely explains the differences from our study findings. Additionally, at our institution, patients are initially tested for a DSA response at 10-14 days post-HTx. Thus, we found a significant increase in dnDSAs three weeks post-HTx, suggesting a memory B-cell response related to PGD. Early inflammatory injury to the donor heart and increased transfusions of blood products^34,35^, both factors associated with PGD, may contribute to allosensitization that leads to dnDSAs. Interestingly, DCD HTx were not associated with dnDSAs in a subgroup analysis, perhaps related to a prior observation that DCD HTx patients experience a different mechanism for PGD with quicker recovery than donation after brain death HTx patients.^36^ As the incidence for PGD continues at a high rate in the contemporary era,^33,37^ earlier DSA testing at 10-14 days post-HTx should be considered in PGD patients and future studies should evaluate specific factors associated with PGD that may cause allosensitization.

### Limitations

This study should be interpreted within the context of several important limitations. First, this was a retrospective study from a single center and carries with it the usual limitations for these studies, including generalizability and the potential confounding of treatment effect. Thus, our findings do not provide guidance for treatment decisions, due to the wide variability in treatment for pAMR in our study. However, only a minority of pAMR+/DSA-patients were treated (23%) and, despite this, the pAMR+/DSA-group had more favorable outcomes when compared to the pAMR+/DSA+ group (73% treated). Second, our center utilizes C4d immunofluorescence with the use of CD68 and C4d immunoperoxidase staining in equivocal cases or when immunofluorescence is not feasible.^1^ However, previous studies have shown that immunofluorescence and immunoperoxidase staining are similarly sensitive and specific for C4d positivity and our prevalence of pAMR was similar to prior studies.^14,38^ Third, we did not evaluate different MFI values for DSAs as the goal of our study was to determine the utility of DSA with pAMR testing using prespecified MFI cutoffs. Additionally, MFI measurements have been known to vary among HLA laboratories, limiting translatability of MFI findings.^39^ Lastly, PGD diagnosis was based on documentation by HTx clinical team members or use of mechanical circulatory support within 24 hours after HTx. Thus, while the PGD rate was similar to most prior studies,^33^ mild and moderate PGD are potentially underestimated, a recognized limitation due to the current PGD criteria.^37^

### Conclusions

Our study findings support the potential use of DSA testing as the primary surveillance method to identify the high-risk pAMR+/DSA+ patients. Additionally, PGD is a novel risk factor for dnDSAs and earlier DSA testing at 10-14 days post-HTx should be considered in patients with PGD.

## Data Availability

All data produced are available online at
https://data.mendeley.com/datasets/d4f7g8hs5z/1

## Non-standard Abbreviations and Acronyms

ACR: Acute cellular rejection
CAV: Cardiac allograft vasculopathy
DCD: Donation after circulatory death
dnDSA: *de novo* donor-specific antibody
DSA: Donor-specific antibody
EMB: Endomyocardial biopsy
HLA: Human leukocyte antigen
HTx: Heart transplantation
ISHLT: International Society for Heart and Lung Transplantation
MFI: Mean fluorescence intensity
pAMR: Pathologic antibody mediated rejection
PGD: Primary graft dysfunction
UCSD: University of California, San Diego Health

## Acknowledgements

The authors acknowledge Priyesha Bijlani, MD, from UCSD for her contribution to data collection and review and editing of prior versions of the article, Hyoungmin Kim, from UCSD for his contribution to data collection, Taylor Tran, BS, from UCSD for his contribution to data collection, Layla Myers, BS, from UCSD for her contribution to data collection, and Michael C. Fishbein, MD, from UCLA for his review and editing of the final manuscript.

---

**Table S1.** Baseline characteristics of heart transplant patients and outcomes for the total study cohort. ACR, acute cellular rejection; AMR, antibody mediated rejection; BMI, body mass index; CMV, cytomegalovirus; DCD, donation after cardiac death; DSA, donor-specific antibodies; HTx, heart transplantation; ICM, ischemic cardiomyopathy; ISHLT, International Society for Heart and Lung Transplantation; MCS, mechanical circulatory support; NICM, nonischemic cardiomyopathy; pAMR, pathologic antibody mediated rejection; PHM, predicted heart mass; PRA, panel reactive antibodies.

| Characteristics | No. of patients<br>(n = 544) | Study cohort |
| --- | --- | --- |
| **Donor characteristics** |  |  |
| Age, y, mean (SD) | 520 | 32.8 (10.6) |
| Male, N (%) | 519 | 428 (82.5) |
| **Recipient characteristics** |  |  |
| Age, y, mean (SD) | 544 | 53.9 (14.3) |
| Male, N (%) | 544 | 436 (80.1) |
| **Race** |  |  |
| Asian, N (%) | 544 | 37 (6.8) |
| Black, N (%) | 544 | 69 (12.7) |
| Native American, N (%) | 544 | 4 (0.7) |
| Other Race, N (%) | 544 | 35 (6.4) |
| Pacific Islander, N (%) | 544 | 12 (2.2) |
| White, N (%) | 544 | 387 (71.1) |
| **Ethnicity** |  |  |
| Hispanic or Latino, N (%) | 544 | 165 (30.3) |
| Recipient BMI, mean (SD) | 521 | 26.5 (4.4) |
| **Indication for HTx** |  |  |
| NICM, N (%) | 544 | 325 (59.7) |
| ICM, N (%) | 544 | 185 (34.0) |
| Congenital, N (%) | 544 | 22 (4.0) |
| Cardiac allograft failure, N (%) | 544 | 12 (2.2) |
| Allosensitization pre-HTx (PRA ≥ 10%), N (%) | 457 | 85 (18.6) |
| Durable MCS, N (%) | 543 | 188 (34.6) |
| **HTx characteristics** |  |  |
| Multiorgan transplant, N (%) | 544 | 76 (14.0) |
| Cold ischemic time, min, mean (SD) | 520 | 200.3 (64.0) |
| Sex mismatch (female D-male R), N (%) | 519 | 42 (8.1) |
| PHM difference, % recipient PHM, mean (SD) | 513 | 5.2 (20.9) |
| Induction therapy, N (%) | 519 | 256 (49.3) |
| DCD, N (%) | 544 | 82 (15.1) |
| CMV mismatch (D+/R-), N (%) | 531 | 105 (19.8) |
| **HTx outcomes** |  |  |
| Primary graft dysfunction | 542 | 118 (21.8) |
| History of pAMR positivity | 544 | 75 (13.8) |
| pAMR1i | 75 | 44 (58.7) |
| pAMR1h | 75 | 5 (6.7) |
| pAMR2 | 75 | 26 (34.7) |
| Mixed ACR (ISHLT grade ≥ 2R) and pAMR | 75 | 8 (10.7) |
| History of DSA positivity | 544 | 140 (25.7) |
| De novo DSA | 134 | 129 (96.3) |
| Class I de novo DSAs alone | 129 | 23 (17.8) |
| Class II de novo DSAs alone | 129 | 75 (58.1) |
| Both class I and II de novo DSAs | 129 | 31 (24.0) |
| Concurrent cardiac allograft<br>vasculopathy with initial pAMR positivity | 20 | 5 (25.0) |
| Concurrent cardiac allograft dysfunction<br>with initial pAMR positivity | 72 | 17 (23.6) |

**Table S2.** Causes of death compared across 1 pAMR/DSA groups. DSA, donor-specific antibody; pAMR, pathologic antibody mediated rejection.

| Outcomes | pAMR+/DSA+<br>(n = 45) | pAMR+/DSA-<br>(n = 30) | pAMR-/DSA+<br>(n = 95) | pAMR-/DSA-<br>(n = 374) | Total |
| --- | --- | --- | --- | --- | --- |
| All-cause mortality or cardiac retransplant (%) | 13 (28.9) | 4 (13.3) | 6 (6.3) | 38 (10.2) | 61 (11.2) |
| Cardiovascular related death (%) | 7 (15.6) | 1 (3.3) | 3 (3.2) | 6 (1.6) | 17 (3.1) |
| Infectious related mortality (%) | 1 (2.2) | 1 (3.3) | 2 (2.1) | 19 (5.1) | 23 (4.2) |
| Cancer (%) | 0 | 0 | 0 | 5 (1.3) | 5 (0.9) |
| Other cause of death (%) | 2 (4.4) | 1 (3.3) | 1 (1.1) | 4 (1.1) | 8 (1.5) |
| Unknown cause of death (%) | 1 (2.2) | 1 (3.3) | 0 | 2 (0.5) | 4 (0.7) |

**Table S3.** Comparison of different de novo DSA patterns for diagnosis of pathologic antibody mediated rejection. DSA, donor-specific antibody. 95% confidence intervals are in parenthesis. #, reference is all other DSAs group.

| De novo DSAs | Positive predictive value | p<sub>c</sub>-value<sup>#</sup> | Odds ratio | p<sub>c</sub>-value<sup>#</sup> |
| --- | --- | --- | --- | --- |
| Both class I and II DSAs on initial DSA+ testing | 64.2% (36.4%-88.9%) | 0.031 | 6.44 (1.14-45.82) | 0.016 |
| Progression from one DSA to both class I and II DSAs | 58.2% (28.6%-87.5%) | 0.124 | 5.02 (0.81-35.88) | 0.087 |
| Two or more class II DSAs on initial DSA+ testing | 52.9% (28.6%-77.3%) | 0.103 | 4.05 (0.85-20.26) | 0.087 |
| All other DSAs | 21.5% (12.9%-30.5%) | - | - | - |

**Table S4.**
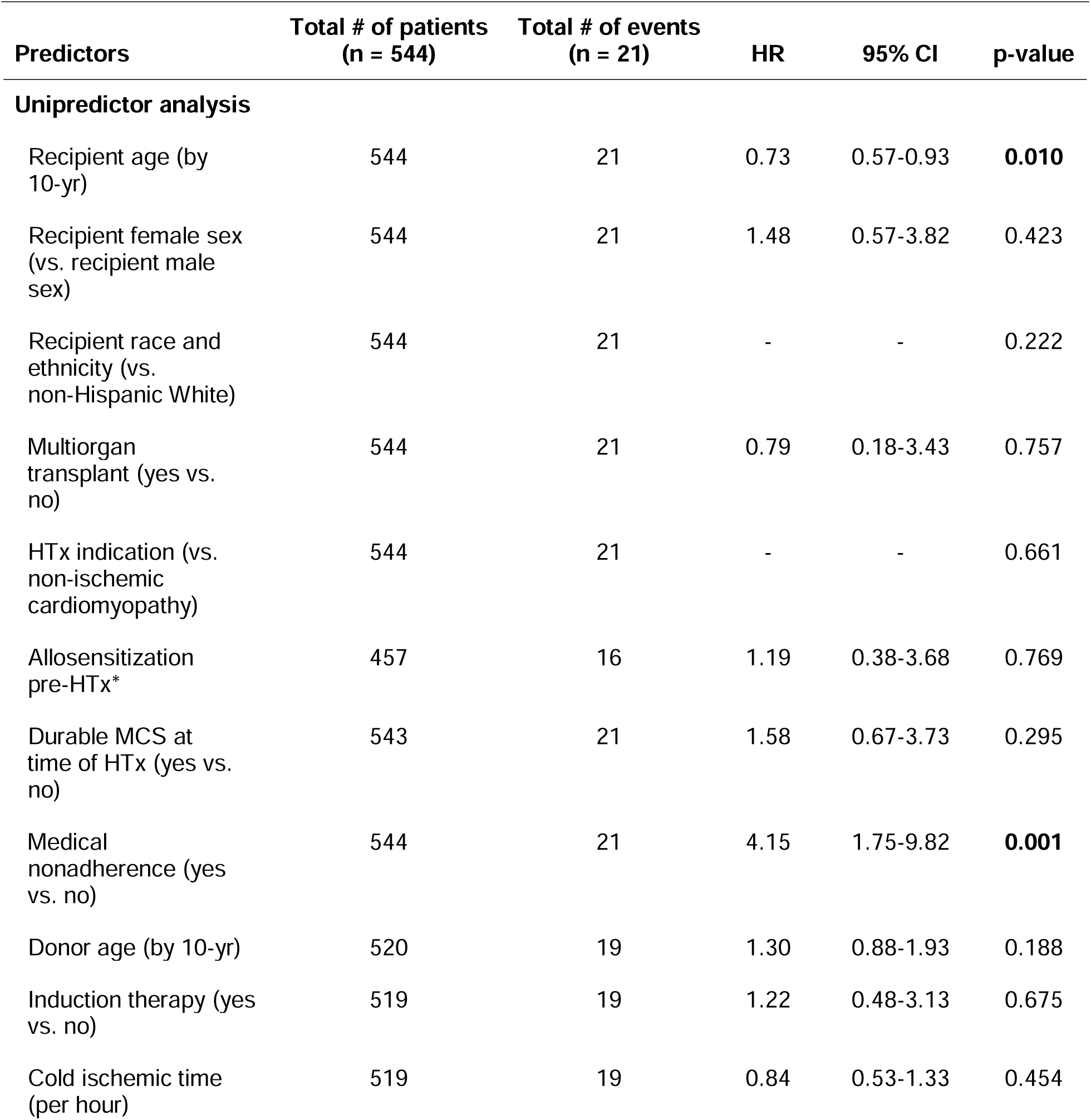
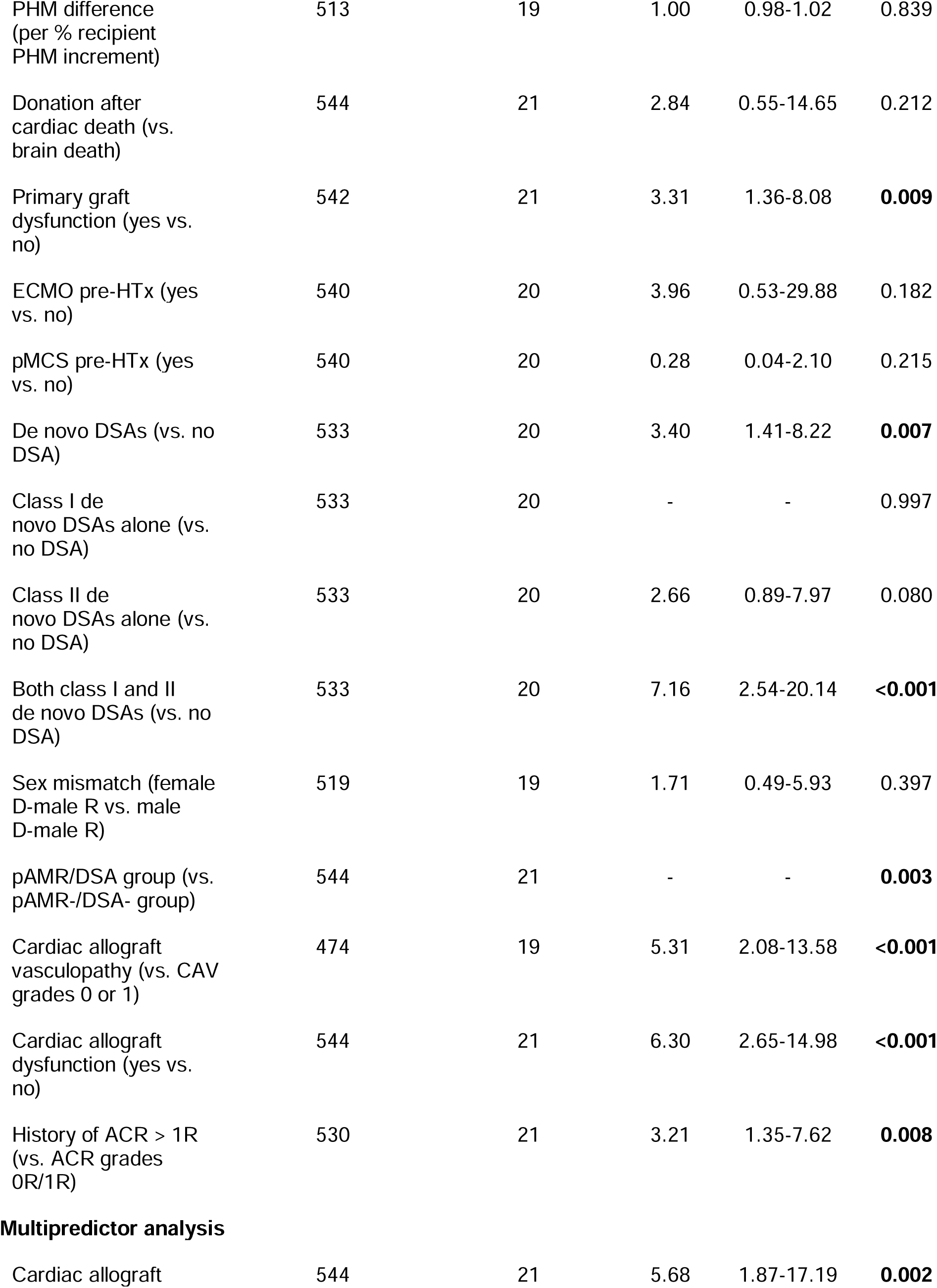
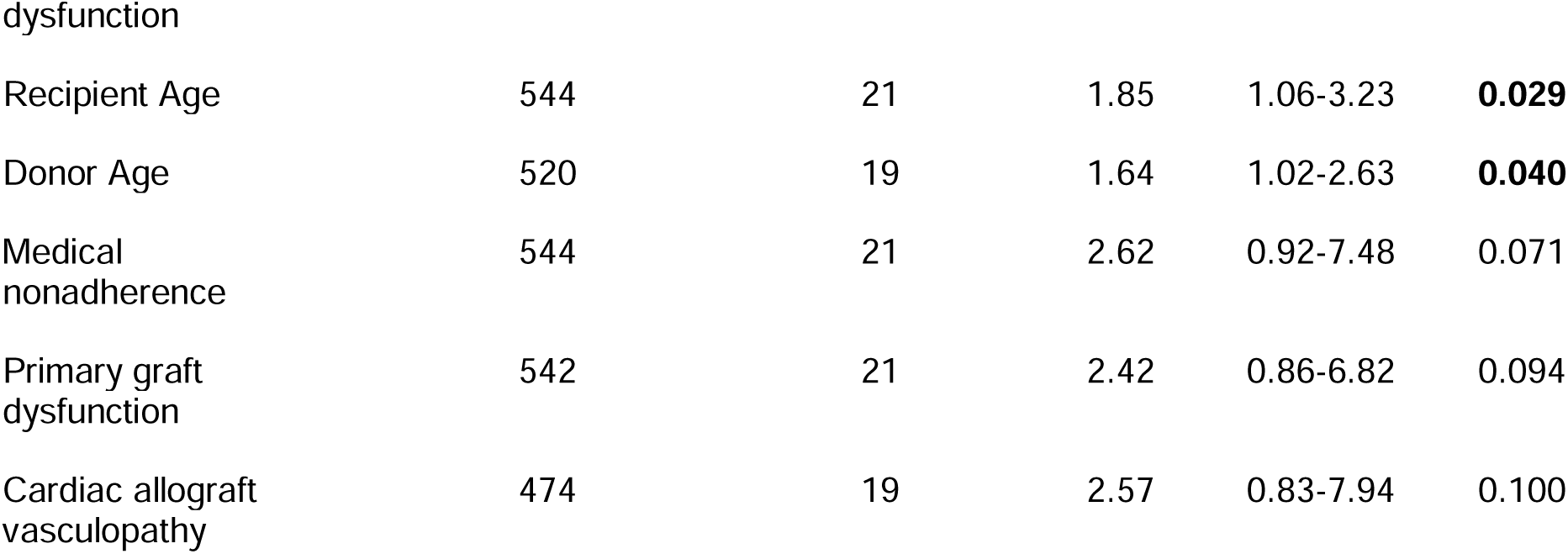
Unipredictor and multipredictor Cox 1 proportional hazards analyses for cardiac survival. Unipredictor parameters with a p-value < 0.15 are included in addition to certain clinical parameters of interest. HRs and CIs are not provided for categorical variables in this table. ACR, acute cellular rejection; CI, confidence interval; cPRA, calculated panel reactive antibodies; DSA, donor-specific antibodies; ECMO, extracorporeal membrane oxygenation; HTx, heart transplantation; HR, hazard ratio; MCS, mechanical circulatory support; pAMR, pathologic antibody mediated rejection; PHM, predicted heart mass; pMCS, percutaneous mechanical circulatory support; UNOS, United Network for Organ Sharing. *, allosensitized patients defined as having a UNOS cPRA >= 10%.

## REFERENCES

1. Berry GJ, Burke MM, Andersen C, et al. The 2013 International Society for Heart and Lung Transplantation Working Formulation for the standardization of nomenclature in the pathologic diagnosis of antibody-mediated rejection in heart transplantation. J Heart Lung Transplant. 2013;32(12):1147–1162. doi:10.1016/j.healun.2013.08.011

2. Stewart S, Winters GL, Fishbein MC, et al. Revision of the 1990 working formulation for the standardization of nomenclature in the diagnosis of heart rejection. J Heart Lung Transplant. 2005;24(11):1710–1720. doi:10.1016/j.healun.2005.03.019

3. Velleca A, Shullo MA, Dhital K, et al. The International Society for Heart and Lung Transplantation (ISHLT) guidelines for the care of heart transplant recipients. J Heart Lung Transplant. 2023;42(5):e1–e141. doi:10.1016/j.healun.2022.10.015

4. Colvin MM, Cook JL, Chang P, et al. Antibody-mediated rejection in cardiac transplantation: emerging knowledge in diagnosis and management: a scientific statement from the American Heart Association. Circulation. 2015;131(18):1608–1639. doi:10.1161/CIR.0000000000000093

5. Khush KK, Hsich E, Potena L, et al. The International Thoracic Organ Transplant Registry of the International Society for Heart and Lung Transplantation: Thirty-eighth adult heart transplantation report - 2021; Focus on recipient characteristics. J Heart Lung Transplant. 2021;40(10):1035–1049. doi:10.1016/j.healun.2021.07.015

6. Cusi V, Vaida F, Wettersten N, et al. Incidence of Acute Rejection Compared With Endomyocardial Biopsy Complications for Heart Transplant Patients in the Contemporary Era. Transplantation. 2024;108(5):1220–1227. doi:10.1097/TP.0000000000004882

7. Zavyalova D, Abraha J, Rao P, Morris GP. Incidence and impact of allele-specific anti-HLA antibodies and high-resolution HLA genotyping on assessing immunologic compatibility. Hum Immunol. 2021;82(3):147–154. doi:10.1016/j.humimm.2021.01.002

8. Coutance G, Kransdorf E, Aubert O, et al. Clinical Prediction Model for Antibody-Mediated Rejection: A Strategy to Minimize Surveillance Endomyocardial Biopsies After Heart Transplantation. Circ Heart Fail. 2022;15(10):e009923. doi:10.1161/CIRCHEARTFAILURE.122.009923

9. Mehra MR, Crespo-Leiro MG, Dipchand A, et al. International Society for Heart and Lung Transplantation working formulation of a standardized nomenclature for cardiac allograft vasculopathy-2010. J Heart Lung Transplant. 2010;29(7):717–727. doi:10.1016/j.healun.2010.05.017

10. Kobashigawa J, Zuckermann A, Macdonald P, et al. Report from a consensus conference on primary graft dysfunction after cardiac transplantation. J Heart Lung Transplant. 2014;33(4):327–340. doi:10.1016/j.healun.2014.02.027

11. Sellarés J, de Freitas DG, Mengel M, et al. Understanding the causes of kidney transplant failure: the dominant role of antibody-mediated rejection and nonadherence. Am J Transplant. 2012;12(2):388–399. doi:10.1111/j.1600-6143.2011.03840.x

12. Kindem IA, Bjerre A, Hammarstrøm C, Naper C, Midtvedt K, Åsberg A. Kidney-transplanted Adolescents—Nonadherence and Graft Outcomes During the Transition Phase: A Nationwide Analysis, 2000–2020. Transplantation. 2023;107(5):1206. doi:10.1097/tp.0000000000004431

13. Wiebe C, Gibson IW, Blydt-Hansen TD, et al. Evolution and clinical pathologic correlations of de novo donor-specific HLA antibody post kidney transplant: Clinical pathologic correlations of DE Novo DSA. Am J Transplant. 2012;12(5):1157–1167. doi:10.1111/j.1600-6143.2012.04013.x

14. Clerkin KJ, Farr MA, Restaino SW, et al. Donor-specific anti-HLA antibodies with antibody-mediated rejection and long-term outcomes following heart transplantation. J Heart Lung Transplant. 2017;36(5):540–545. doi:10.1016/j.healun.2016.10.016

15. Smith JD, Banner NR, Hamour IM, et al. De novo donor HLA-specific antibodies after heart transplantation are an independent predictor of poor patient survival. Am J Transplant. 2011;11(2):312–319. doi:10.1111/j.1600-6143.2010.03383.x

16. Setia G, Nishihara K, Singer Englar T, Zhang X, Patel J, Kobashigawa J. Crossing low/moderate level donor specific antibodies during heart transplantation. Clinical Transplantation. 2021;35(3). doi:10.1111/ctr.14196

17. Agbor-Enoh S, Shah P, Tunc I, et al. Cell-Free DNA to Detect Heart Allograft Acute Rejection. Circulation. 2021;143(12):1184–1197. doi:10.1161/CIRCULATIONAHA.120.049098

18. Baldwin WM 3rd, Samaniego-Picota M, Kasper EK, et al. Complement deposition in early cardiac transplant biopsies is associated with ischemic injury and subsequent rejection episodes. Transplantation. 1999;68(6):894–900. doi:10.1097/00007890-199909270-00024

19. Collard CD, Väkevä A, Morrissey MA, et al. Complement activation after oxidative stress: role of the lectin complement pathway. Am J Pathol. 2000;156(5):1549–1556. doi:10.1016/S0002-9440(10)65026-2

20. Mantell BS, Cordero H, See SB, et al. Transcriptomic heterogeneity of antibody mediated rejection after heart transplant with or without donor specific antibodies. J Heart Lung Transplant. 2021;40(11):1472–1480. doi:10.1016/j.healun.2021.06.012

21. Butler CL, Hickey MJ, Jiang N, et al. Discovery of non-HLA antibodies associated with cardiac allograft rejection and development and validation of a non-HLA antigen multiplex panel: From bench to bedside. Am J Transplant. 2020;20(10):2768–2780. doi:10.1111/ajt.15863

22. Reed EF, Demetris AJ, Hammond E, et al. Acute antibody-mediated rejection of cardiac transplants. J Heart Lung Transplant. 2006;25(2):153–159. doi:10.1016/j.healun.2005.09.003

23. Clerkin KJ, Restaino SW, Zorn E, Vasilescu ER, Marboe CC, Mancini DM. The effect of timing and graft dysfunction on survival and cardiac allograft vasculopathy in antibody-mediated rejection. J Heart Lung Transplant. 2016;35(9):1059–1066. doi:10.1016/j.healun.2016.04.007

24. Tan CD, Sokos GG, Pidwell DJ, et al. Correlation of Donor-Specific Antibodies, Complement and Its Regulators with Graft Dysfunction in Cardiac Antibody-Mediated Rejection. American Journal of Transplantation. 2009;9(9):2075–2084. doi:10.1111/j.1600-6143.2009.02748.x

25. Hodges AM, Lyster H, McDermott A, et al. Late antibody-mediated rejection after heart transplantation following the development of de novo donor-specific human leukocyte antigen antibody. Transplantation. 2012;93(6):650–656. doi:10.1097/TP.0b013e318244f7b8

26. Pham MX, Teuteberg JJ, Kfoury AG, et al. Gene-expression profiling for rejection surveillance after cardiac transplantation. N Engl J Med. 2010;362(20):1890–1900. doi:10.1056/NEJMoa0912965

27. Zhang X, Kransdorf E, Levine R, Patel JK, Kobashigawa JA. HLA-DQ mismatches stimulate de novo donor specific antibodies in heart transplant recipients. Hum Immunol. 2020;81(7):330–336. doi:10.1016/j.humimm.2020.04.003

28. DeVos JM, Gaber AO, Knight RJ, et al. Donor-specific HLA-DQ antibodies may contribute to poor graft outcome after renal transplantation. Kidney Int. 2012;82(5):598–604. doi:10.1038/ki.2012.190

29. Ho EK, Vasilescu ER, Vlad G, Clynes RA, Ratner LE, Suciu-Foca N. Detection of donor-specific-antibodies by solid phase assay and its relevance to complement-dependent-lymphocytotoxicity cross-matching in kidney transplantation. Hum Immunol. 2014;75(6):520–523. doi:10.1016/j.humimm.2014.03.002

30. Irving CA, Carter V, Gennery AR, et al. Effect of persistent versus transient donor-specific HLA antibodies on graft outcomes in pediatric cardiac transplantation. J Heart Lung Transplant. 2015;34(10):1310–1317. doi:10.1016/j.healun.2015.05.001

31. Cross AR, Lion J, Poussin K, et al. HLA-DQ alloantibodies directly activate the endothelium and compromise differentiation of FoxP3high regulatory T lymphocytes. Kidney Int. 2019;96(3):689–698. doi:10.1016/j.kint.2019.04.023

32. Han J, Moayedi Y, Henricksen EJ, et al. Primary Graft Dysfunction Is Associated With Development of Early Cardiac Allograft Vasculopathy, but Not Other Immune-mediated Complications, After Heart Transplantation. Transplantation. 2023;107(7):1624–1629. doi:10.1097/TP.0000000000004551

33. Benck L, Kransdorf EP, Emerson DA, et al. Recipient and surgical factors trigger severe primary graft dysfunction after heart transplant. J Heart Lung Transplant. 2021;40(9):970–980. doi:10.1016/j.healun.2021.06.002

34. McDonald MM, Mihalj M, Zhao B, et al. Clinicopathological correlations in heart transplantation recipients complicated by death or re-transplantation. Front Cardiovasc Med. 2022;9. doi:10.3389/fcvm.2022.1014796

35. Holmström EJ, Syrjälä SO, Dhaygude K, et al. Severe primary graft dysfunction of the heart transplant is associated with increased plasma and intragraft proinflammatory cytokine expression. J Heart Lung Transplant. 2023;42(6):807–818. doi:10.1016/j.healun.2023.01.005

36. Ayer A, Truby LK, Schroder JN, et al. Improved outcomes in severe primary graft dysfunction after heart transplantation following donation after circulatory death compared with donation after brain death. J Card Fail. 2023;29(1):67–75. doi:10.1016/j.cardfail.2022.10.429

37. Truby LK, Takeda K, Topkara VK, et al. Risk of severe primary graft dysfunction in patients bridged to heart transplantation with continuous-flow left ventricular assist devices. J Heart Lung Transplant. 2018;37(12):1433–1442. doi:10.1016/j.healun.2018.07.013

38. Chantranuwat C, Qiao JH, Kobashigawa J, Hong L, Shintaku P, Fishbein MC. Immunoperoxidase staining for C4d on paraffin-embedded tissue in cardiac allograft endomyocardial biopsies: comparison to frozen tissue immunofluorescence. Appl Immunohistochem Mol Morphol. 2004;12(2):166–171. doi:10.1097/00129039-200406000-00012

39. Reed EF, Rao P, Zhang Z, et al. Comprehensive assessment and standardization of solid phase multiplex-bead arrays for the detection of antibodies to HLA. Am J Transplant. 2013;13(7):1859–1870. doi:10.1111/ajt.12287

